# Characterizing hepatitis B virus infection in children in the Democratic Republic of Congo to inform elimination efforts

**DOI:** 10.1101/2024.06.12.24308840

**Authors:** CE Morgan, KA Powers, JK Edwards, U Devkota, S Biju, FC Lin, JL Schmitz, G Cloherty, J Muwonga, A Mboyo, P Tshiamala, MM Kashamuka, A Tshefu, M Emch, M Yotebieng, S Becker-Dreps, JB Parr, P Thompson

## Abstract

**Objective:** Despite global reductions in hepatitis B virus (HBV) prevalence, an estimated 6.2 million children are infected, two-thirds of whom live in the WHO Africa region. We sought to characterize childhood HBV to inform elimination efforts in the Democratic Republic of Congo (DRC), one of the largest and most populous African countries.

**Methods:** Using the most recent (2013–14) nationally representative Demographic and Health Survey in the DRC, we analyzed HBV surface antigen (HBsAg) on dried blood spots and associated survey data from children aged 6-59 months. We estimated HBsAg-positivity prevalence nationally, regionally, and by potential correlates of infection. We evaluated spatial variation in HBsAg-positivity prevalence, overall and by age, sex, and vaccination status.

**Findings:** Using data from 5,679 children, we found national HBsAg-positivity prevalence was 1.3% (95% CI: 0.9%-1.7%), but ranged from 0.0% in DRC’s capital city province, Kinshasa, to 5.6% in northwestern Sud-Ubangi Province. Prevalence among boys (1.8%, 95% CI: 1.2%-2.7%) was double that among girls (0.7%, 95%CI: 0.4%-1.3%). Tetanus antibody-negativity, rurality, and lower household wealth were also significantly associated with higher HBsAg-positivity prevalence. We observed no difference in prevalence by age. Children had higher HBsAg-positivity odds if living with ≥1 HBsAg-positive adult household member (OR: 2.3, 95%CI: 0.7-7.8), particularly an HBsAg-positive mother (OR: 7.2, 95%CI:1.6-32.2).

**Conclusion:** In the largest national survey of HBV among children and household contacts in the DRC, we found that childhood HBV prevalence was 10-60 times the global target of 0.1%. We highlight specific regions and populations for further investigation and focused prevention efforts.

## Introduction

An estimated 6.2 million children are infected with hepatitis B virus (HBV) globally, two-thirds of whom live in Africa.(1) Prevention of HBV transmission in children is critical, as they have a substantially higher risk of developing chronic HBV infection following exposure (>90% in infancy) compared with adults (<10%).(2) HBV prevention in children is focused on prevention of perinatal transmission, with HBsAg testing during pregnancy, antenatal antiviral prophylaxis administered to mothers with high serum HBV DNA levels (≥200,000 IU/mL), HBV birth-dose vaccination, and post-exposure prophylaxis with HBV immunoglobulin where available, along with a three-dose vaccine series (HepB3) given in infancy.(3) While these interventions are highly efficacious and acceptable in preventing perinatal transmission,(4–6) they are not fully implemented in many parts of Africa.(7) Further, epidemiological studies from several African countries suggest that early horizontal transmission may contribute more to high HBV prevalence in children than in other regions globally,(8–12) but the viral, immunological, and sociocultural causes of these transmission patterns remain poorly understood, such as in the Democratic Republic of Congo (DRC), one of the 2024 WHO viral hepatitis focus countries.

HBV prevalence remains poorly defined in the DRC, which is the fourth most populous African country, has the seventh-highest birth rate, and is the second largest country on the continent by land mass.(13,14) The estimates that are available for central Africa broadly suggest an HBsAg-positivity prevalence of 10%-14% and that fewer than 1% of infections are diagnosed.(15) Due to the lack of nationally representative data in the region, central African countries contribute the largest uncertainty to global estimates.(16)

We previously analyzed dried blood spot (DBS) samples from 277 children across the DRC, finding an HBV prevalence of 2.2%,(17) but evaluation of spatial variation and infection correlates was limited by sample size. In the current study, we analyzed all remaining samples (>5,600) of children aged 6-59 months from the most recent (2013–14) Demographic and Health Survey (DHS) in the DRC to characterize HBV epidemiology and inform HBV elimination efforts in this setting. Collected six years after HepB3 was introduced in the national infant immunization program,(18,19) these data enable a preliminary assessment of HepB3 impact through evaluation of HBV prevalence by age in children <5 years old. We further evaluate HBV prevalence within subgroups of interest, including by sex and province, with the goal of identifying populations for focused interventions.

## Methods

### Study population and design

We characterized HBV epidemiology among children 6-59 months through three primary objectives: estimation of childhood HBsAg-positivity prevalence nationally, provincially, and by subgroup, including by age group and sex; assessment of correlates of HBsAg-positivity prevalence among children; and estimation of odds of infection among children given exposure to an HBV-infected adult household member through a nested case-control study. We used survey data and DBS specimens from the 2013-14 DRC DHS, a population-representative survey that included DBS collection in a 50% random sample of households (**Supplementary Material**). Detailed survey methodology is described in the 2013-2014 DRC DHS final report.(20)

For the first and second study objectives, all children with sufficient DBS sample available were included. For the nested case-control study in the third objective, case households were defined as those with exactly one child aged 6-59 months who was HBsAg-positive in our biospecimen analysis (described below); these children (no more than one per household) formed the case population. Control households were sampled in a 6:1 ratio (control:case households) from households in which all children tested were HBsAg-negative; these children (possibly more than one per household) were the controls. Within these case and control households, adult household members with DBS collected were evaluated for HBV infection as a potential correlate of infection for the case vs. control children.

### Biospecimen analysis

We determined HBV status by eluting DBS samples and analyzing HBsAg presence on the ARCHITECT platform (Abbott Laboratories, Abbot Park, IL, USA)(21) in a CLIA-approved laboratory (**Supplementary Material**). The ARCHITECT instrument defines a positive HBsAg result using a ratio of “signal-to-cutoff” (S/CO). To reduce the risk of false-positive results, we employed a two-step approach in interpreting a positive HBsAg result. First, any sample with S/CO >1 was retested automatically using the same eluant (technical replicate). Second, if there was sufficient DBS sample remaining, the assay was repeated using a fresh 6mm DBS punch (biological replicate) for any positive sample with a S/CO between 1-100 or with an adjacent positive on the elution plate. Samples with insufficient material for repeat testing were considered positive in the primary analysis if the average S/CO of both technical replicates was ≥5, and negative otherwise. All other samples were considered positive if both positive biological replicates (four total technical replicates) were positive (S/CO ≥5), and negative otherwise. We conducted sensitivity analyses with different positivity cut-offs (S/CO of 1, 2, and 100).

### HBsAg prevalence and correlates analysis

For representative estimates of HBsAg positivity prevalence, we applied two types of weights. To account for the probability of selection into the survey, we used DHS survey weights available through the DHS Program. To account for sample missingness or exhaustion among those selected to provide biospecimens, we followed an established approach to calculate propensity score weights using inverse probability of sampling (elsewhere called “treatment”) weighting.(22,23) Briefly, we identified attributes that could be associated with the outcome (HBsAg status) or sample missingness/exhaustion (**Supplementary Material**). These variables were included as independent predictors in a binomial logistic regression model in which having an HBsAg result (yes/no) was the dependent variable. We calculated predicted probabilities from this model to obtain propensity scores, the inverse of which formed the weights. We stabilized the inverse propensity score weights by dividing each weight by the sum of weights in the group with samples. The stabilized weights were multiplied by the DHS survey weights to create final survey weights in the analysis.

We identified variables available from the DHS survey to assess as potential correlates of HBV infection. Details on variable coding are available in Supplementary Material, but briefly: variables included were age, sex, province, rurality, location (provincial capital, small city, town, countryside), household wealth, anemia, *P. falciparum* malaria infection, tetanus serology results (a proxy for pentavalent vaccination, which includes diphtheria-pertussis-tetanus [DPT] and HepB3), reported number of DPT vaccine doses, growth stunting, injections received in the last 12 months (such as antibiotics and antimalarial medications), use of a new and unopened needle/syringe in the last injection, and reporting justification of violence toward women in the household. *P. falciparum* infection was determined using quantitative real-time PCR targeting the *P. falciparum* lactate dehydrogenase (*pfldh*) gene, conducted prior to this study and previously described.(24)*Plasmodium* and HBV both infect hepatocytes, but the biological mechanisms of coinfection are poorly characterized. Malaria is known to dampen immunologic responses to polysaccharide and possibly protein-based vaccines,(25) and frequent malaria infections, common in high-prevalence areas like the DRC, result in a higher frequency of blood transfusions, a known risk factor for HBV infection.(26–28) Tetanus antibody results are available publicly through the DHS Program and were produced using an ELISA-based assay.(29)

We described the distribution of the overall population according to each potential correlate of interest and estimated HBsAg-positivity prevalence and Wald-type 95% confidence intervals (CI) within strata of each. We further estimated prevalence differences and associated Wald-type 95% CIs in HBsAg-positivity across correlate strata.

### Spatial analysis

We characterized spatial variation in HBsAg-positivity at the DHS cluster level and at the province level. GPS locations were available through the DHS Program for DHS clusters; for clusters with missing GPS location, we used province designation to randomly select latitude and longitude coordinates within the province to impute GPS coordinates. We calculated prevalence of HBsAg-positivity within each DHS cluster. We also analyzed choropleth maps of weighted HBsAg-positivity prevalence at the province level, overall and by age (categorizing age in months according to age in years), sex (boys vs girls), and HepB3 vaccination proxies (tetanus antibody-positive versus antibody-negative and reported receipt of any DPT vaccine dose versus none). To inform focused public health action, we mapped province-level prevalence differences by these same variables, which we chose for province-level analysis on the basis of expected regional differences in vaccine distribution efforts and cultural practices that could confer differential HBV exposure by sex.

### Nested case-control analysis

For the nested case-control study, we used logistic regression to calculate weighted, unadjusted odds ratios comparing HBsAg positivity in case vs control children according to two separate exposure variables: 1) exposure to at least one HBsAg-positive adult household member of any type, and 2) exposure to an HBsAg-positive mother, specifically.

All data were imported and analyzed in R (v4.3.3, R Core Team, Vienna, Austria),(30) using the *tidyverse* (v2.0.0), *survey* (v4.1-1), and *srvr* (v1.2.0) packages. Spatial analysis was conducted using the *sf* (v.1.0-11) package. R code is publicly available at https://github.com/IDEELResearch/dhs_hbv. This study was approved by the Institutional Review Board at UNC and the Ethics Committee at Kinshasa School of Public Health.

## Results

### Study population

We analyzed 5,679 samples from children 6-59 months of age (**Figure 1**). The mean age among children was 32 months (SD 0.23), and their households had an average of seven residents (**Table 1**). Most children (70%) were from rural areas, and 45% resided in households in the lower two wealth quintiles (as defined by the DHS Program for the full DRC population). Similar percentages of boys and girls were studied. Twenty-eight percent had PCR-confirmed *P. falciparum* infections, and about a third (36%) had detectable tetanus antibodies. Nutritional status was poor overall: almost half (44%) were moderately-to-severely stunted and about a third (35%) had moderate-to-severe anemia. About half of children (53%) had documentation of DPT vaccine completion, and 18% had not initiated the series. Almost one-third of children (31%) had received at least one injection in the last year, and for 7% of children, a reused needle or syringe was applied in their last injection. Approximately two-thirds of children (69%) lived in a household in which physical violence toward a woman was reported as justified. These distributions of demographics of children evaluated for HBV were similar to those of all children selected for DHS biospecimen collection (**Supplementary Table 1**).

**Figure 1.**
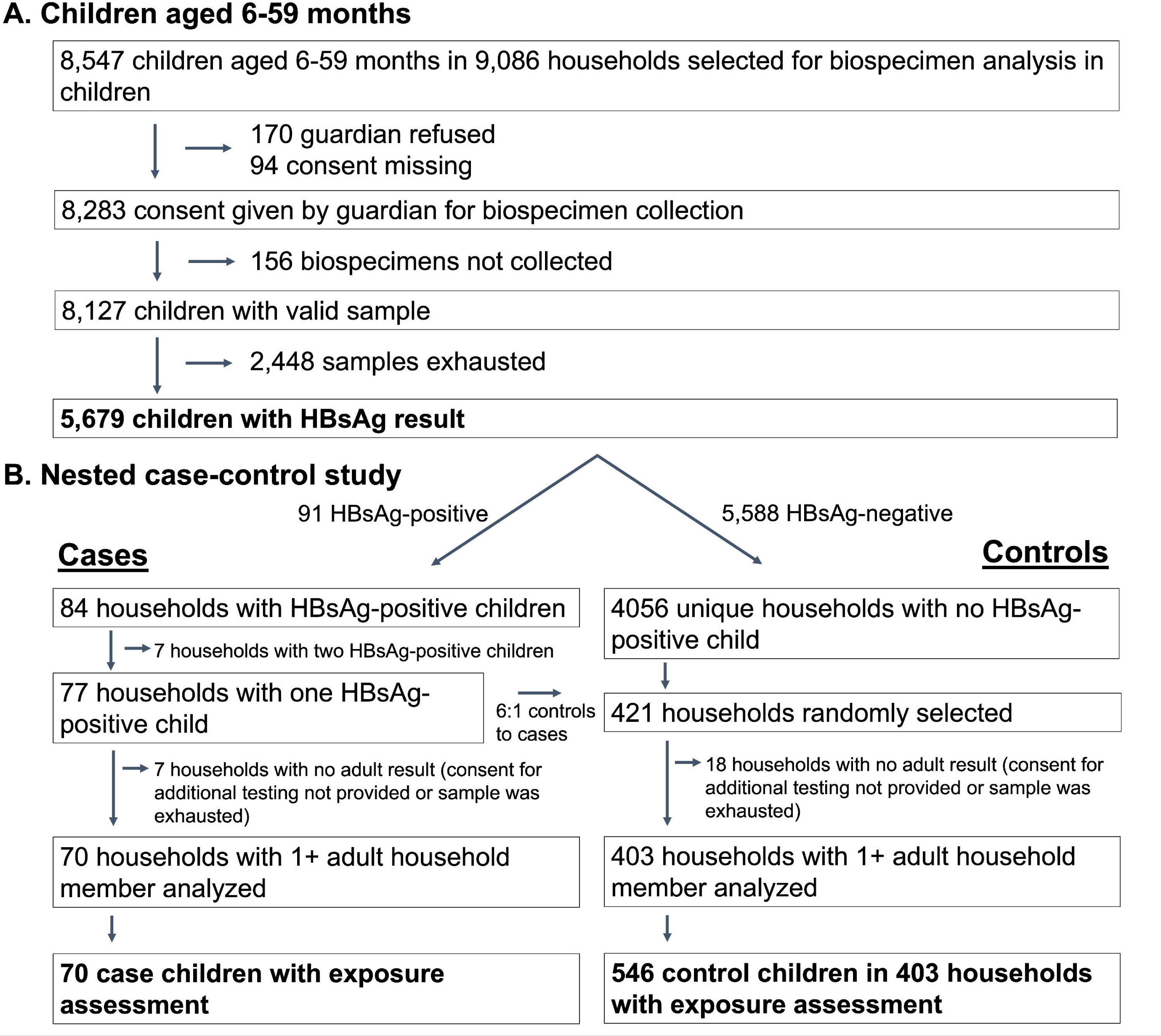
Flowchart of study participants.

**Table 1.** Characteristics of the study population, children 6-59 months.

|  | **Overall\***<br>(N = 5773) | **Nested case-control study** |  |
| --- | --- | --- | --- |
|  |  | **Cases\***<br>(N = 67) | **Controls\***<br>(N=592) |
| **Age (months), mean (SD)** | 32.3 (0.23) | 29.9 (0.82) | 31.9 (2.67) |
| **Household size, mean (SD)** | 6.9 (0.09) | 7.1 (0.24) | 6.5 (0.4) |
| **Number of children <5, mean (SD)** | 2.2 (0.03) | 2.3 (0.09) | 2.3 (0.22) |
| **Age (months), n (%)** |  |  |  |
| 6-11 | 683 (12) | 8 (12) | 92 (16) |
| 12-23 | 1235 (21) | 12 (18) | 133 (22) |
| 24-25 | 1297 (22) | 16 (24) | 138 (23) |
| 36-47 | 1260 (22) | 18 (28) | 128 (22) |
| 48-59 | 1298 (22) | 13 (19) | 101 (17) |
| **Male sex, n (%)** | 2917 (51) | 49 (73) | 302 (51) |
| **Relationship to head of household** |  |  |  |
| Child | 4520 (78) | 54 (81) | 465 (79) |
| Grandchild | 901 (16) | 8 (12) | 85 (14) |
| Other | 352 (6) | 5 (7) | 41 (7) |
| **Rural location, n (%)** | 4053 (70) | 55 (82) | 421 (71) |
| **Location of residence, n (%)** |  |  |  |
| Countryside | 4053 (70) | 55 (82) | 421 (71) |
| Provincial capital | 958 (17) | 8 (12) | 111 (19) |
| Town | 623 (11) | 4 (6) | 50 (8) |
| Small city | 139 (2) | 0 (0) | 10 (2) |
| **Wealth, n (%)** |  |  |  |
| Poorest | 1310 (23) | 23 (34) | 119 (20) |
| Poorer | 1260 (22) | 19 (28) | 130 (22) |
| Middle | 1159 (20) | 13 (19) | 109 (18) |
| Richer | 1149 (20) | 9 (13) | 148 (25) |
| Richest | 895 (15) | 4 (7) | 86 (14) |
| ***P. falciparum* infection<sup>†</sup>, n (%)** | 1629 (28) | 22 (33) | 132 (22) |
| **Moderate-to-severe stunting, n (%)** | 2564 (44) | 32 (48) | 276 (47) |
| **Moderate-to-severe anemia, n (%)** | 1994 (35) | 28 (42) | 190 (32) |
| **Tetanus antibody-positive<sup>‡</sup>, n (%)** | 2092 (36) | 14 (21) | 261 (44) |
| **Diphtheria-pertussis-tetanus vaccination<sup>¶</sup>, n (%)** |  |  |  |
| Series completed | 3057 (53) | 28 (42) | 355 (60) |
| Series incomplete | 949 (16) | 5 (8) | 92 (16) |
| No doses received | 1034 (18) | 21 (31) | 84 (14) |
| Not available | 732 (13) | 13 (19) | 61 (10) |
| **Injectable medications in last year, n (%)** |  |  |  |
| 1-12 | 1436 (25) | 13 (20) | 192 (33) |
| 13-24 | 222 (4) | 2 (4) | 10 (2) |
| ≥25 | 141 (2) | 3 (4) | 11 (2) |
| **None** | 3254 (56) | 36 (54) | 318 (54) |
| **Not available** | 719 (12) | 13 (19) | 60 (10) |
| **Reuse of needles/syringes#, n (%)** |  |  |  |
| **New, unopened syringe/needle applied** | 1647 (93) | 15 (84) | 197 (93) |
| **Used/opened syringe/needle applied** | 125 (7) | 3 (16) | 13 (6) |
| **Physical violence toward wife reported to be justified in household\*\*, n (%)** | 3955 (69) | 47 (70) | 425 (72) |

\* Weighted by DHS sampling weights and inverse propensity score weights accounting for sample missingness/exhaustion.
† *Plasmodium falciparum* infection determined by PCR targeting the *PfIdh* gene, previously described(24).
‡ Tetanus serology available through DHS Programme, and previously described by Cheng *et al.* (2022)(29).
¶ As of 2007, the DPT series included hepatitis B vaccination.

### Prevalence of HBsAg positivity, overall and by correlates of infection

Overall HBsAg-positivity prevalence in children 6-59 months was 1.3% (95% CI: 0.9%, 1.7%) (**Table 2**), ranging from 1.2% to 1.7% in sensitivity analyses (**Supplementary Table 2**). HBsAg-positivity prevalence was 1.8% (95% CI: 1.2%, 2.7%) in boys and 0.7% in girls (95% CI: 0.4%, 1.3%), corresponding to a prevalence difference (PD) of 1.1 cases (95% CI: 0.2-2.0) per 100 children (**Figure 2**). HBsAg-positivity prevalence did not differ appreciably by age, ranging from 1.0% in 12–to-23-month-olds to 1.6% among 36-to-47-month-olds (**Table 2**). Prevalence among children in rural areas (1.5%, 95% CI: 1.0%, 2.1%) was nearly double that of children from urban areas (0.8%, 95% CI: 0.4%, 1.5%). Prevalence decreased with increasing household wealth, from 1.9% (95% CI: 1.1%, 3.1%) in the poorest wealth quintile to 0.6% (95% CI: 0.2%, 2.1%) in the richest wealth quintile. HBsAg-positivity prevalence among children infected with *P. falciparum* was 1.5% (95% CI: 1.0%, 2.2%), compared to 1.2% (95% CI: 0.8%, 1.8%) among those uninfected with *P. falciparum*.

**Figure 2.**
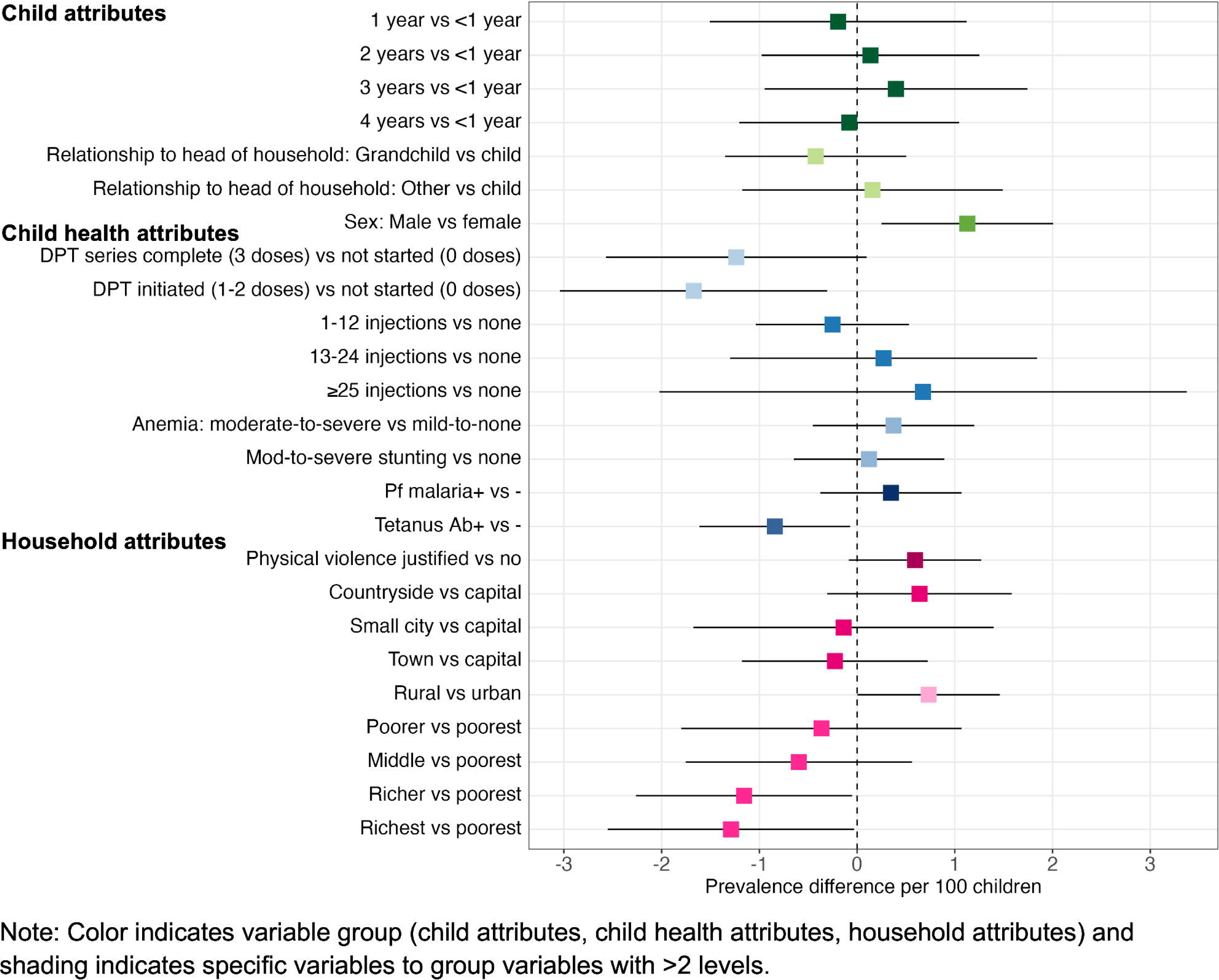
HBsAg-positivity prevalence differences for potential correlates of interest among children 6-59 months.

**Table 2.**
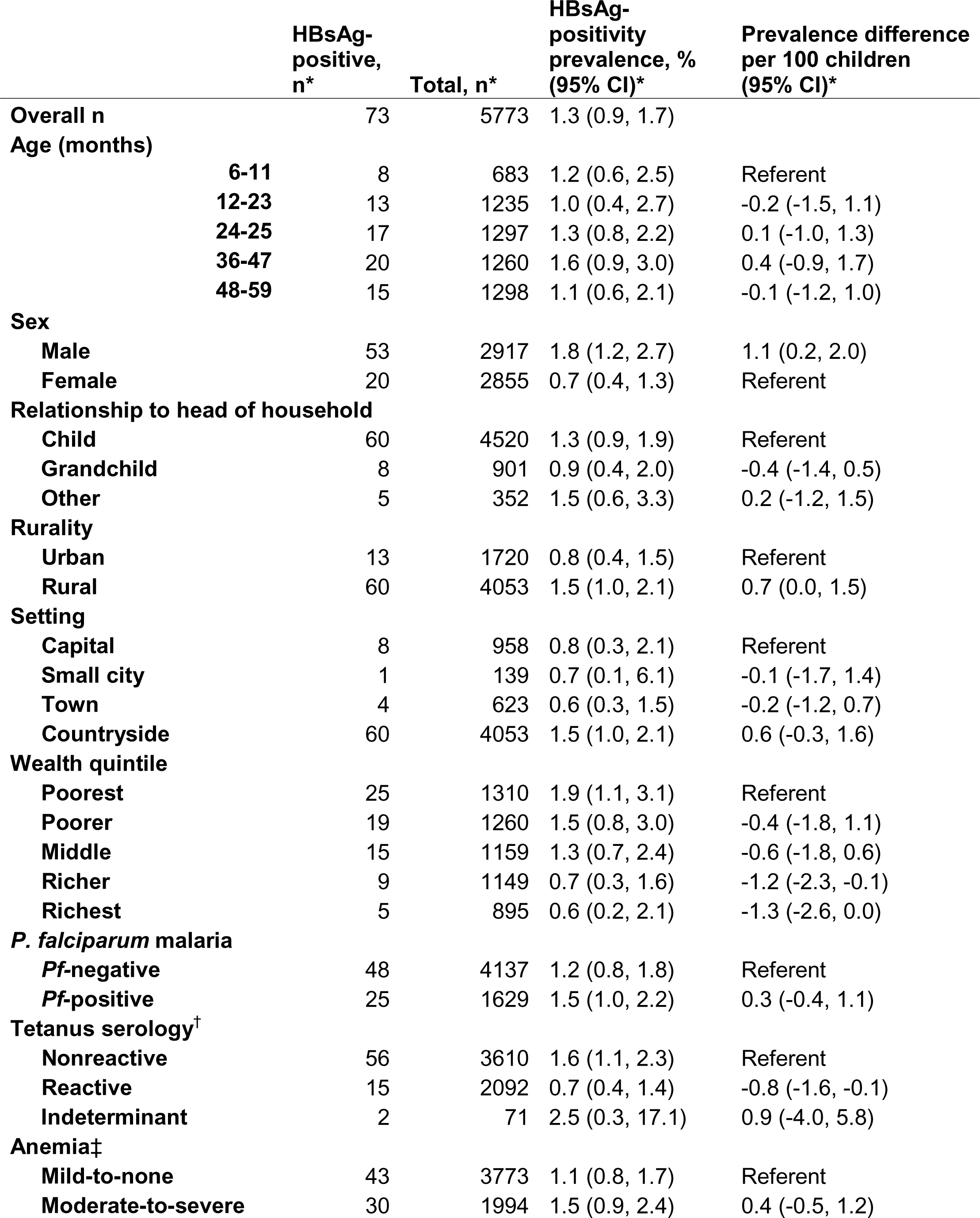
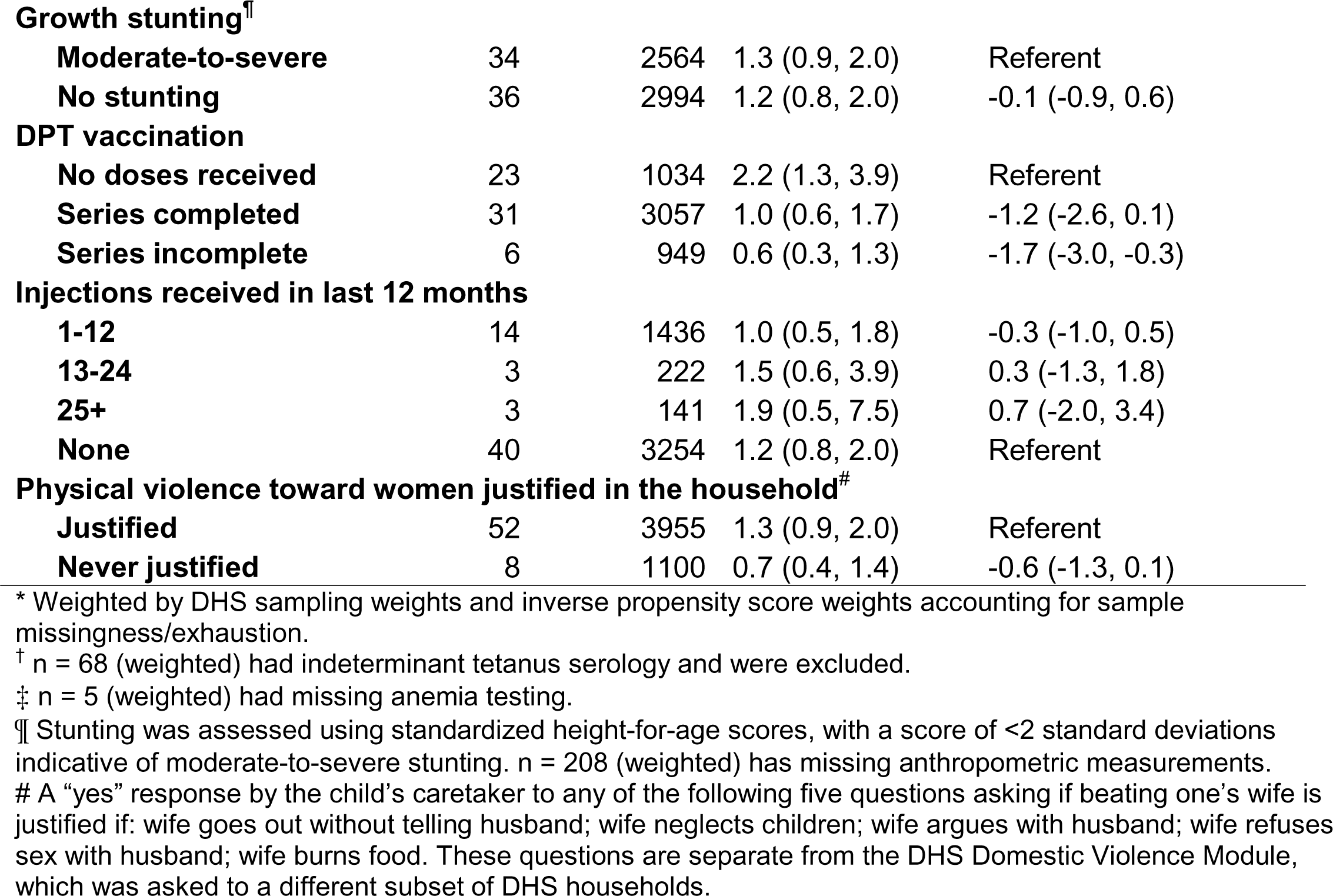
HBsAg-positivity prevalence for children 6-59 months, overall and by potential correlates of interest.

|  | HBsAg-positive,<br>n* | Total, n* | HBsAg-positivity<br>prevalence, %<br>(95% CI)* | Prevalence difference<br>per 100 children<br>(95% CI)* |
| --- | --- | --- | --- | --- |
| **Overall n** | 73 | 5773 | 1.3 (0.9, 1.7) |  |
| **Age (months)** |  |  |  |  |
| **6-11** | 8 | 683 | 1.2 (0.6, 2.5) | Referent |
| **12-23** | 13 | 1235 | 1.0 (0.4, 2.7) | -0.2 (-1.5, 1.1) |
| **24-25** | 17 | 1297 | 1.3 (0.8, 2.2) | 0.1 (-1.0, 1.3) |
| **36-47** | 20 | 1260 | 1.6 (0.9, 3.0) | 0.4 (-0.9, 1.7) |
| **48-59** | 15 | 1298 | 1.1 (0.6, 2.1) | -0.1 (-1.2, 1.0) |
| **Sex** |  |  |  |  |
| **Male** | 53 | 2917 | 1.8 (1.2, 2.7) | 1.1 (0.2, 2.0) |
| **Female** | 20 | 2855 | 0.7 (0.4, 1.3) | Referent |
| **Relationship to head of household** |  |  |  |  |
| **Child** | 60 | 4520 | 1.3 (0.9, 1.9) | Referent |
| **Grandchild** | 8 | 901 | 0.9 (0.4, 2.0) | -0.4 (-1.4, 0.5) |
| **Other** | 5 | 352 | 1.5 (0.6, 3.3) | 0.2 (-1.2, 1.5) |
| **Rurality** |  |  |  |  |
| **Urban** | 13 | 1720 | 0.8 (0.4, 1.5) | Referent |
| **Rural** | 60 | 4053 | 1.5 (1.0, 2.1) | 0.7 (0.0, 1.5) |
| **Setting** |  |  |  |  |
| **Capital** | 8 | 958 | 0.8 (0.3, 2.1) | Referent |
| **Small city** | 1 | 139 | 0.7 (0.1, 6.1) | -0.1 (-1.7, 1.4) |
| **Town** | 4 | 623 | 0.6 (0.3, 1.5) | -0.2 (-1.2, 0.7) |
| **Countryside** | 60 | 4053 | 1.5 (1.0, 2.1) | 0.6 (-0.3, 1.6) |
| **Wealth quintile** |  |  |  |  |
| **Poorest** | 25 | 1310 | 1.9 (1.1, 3.1) | Referent |
| **Poorer** | 19 | 1260 | 1.5 (0.8, 3.0) | -0.4 (-1.8, 1.1) |
| **Middle** | 15 | 1159 | 1.3 (0.7, 2.4) | -0.6 (-1.8, 0.6) |
| **Richer** | 9 | 1149 | 0.7 (0.3, 1.6) | -1.2 (-2.3, -0.1) |
| **Richest** | 5 | 895 | 0.6 (0.2, 2.1) | -1.3 (-2.6, 0.0) |
| ***P. falciparum* malaria** |  |  |  |  |
| ***Pf*-negative** | 48 | 4137 | 1.2 (0.8, 1.8) | Referent |
| ***Pf*-positive** | 25 | 1629 | 1.5 (1.0, 2.2) | 0.3 (-0.4, 1.1) |
| **Tetanus serology<sup>†</sup>** |  |  |  |  |
| **Nonreactive** | 56 | 3610 | 1.6 (1.1, 2.3) | Referent |
| **Reactive** | 15 | 2092 | 0.7 (0.4, 1.4) | -0.8 (-1.6, -0.1) |
| **Indeterminant** | 2 | 71 | 2.5 (0.3, 17.1) | 0.9 (-4.0, 5.8) |
| **Anemia<sup>‡</sup>** |  |  |  |  |
| **Mild-to-none** | 43 | 3773 | 1.1 (0.8, 1.7) | Referent |
| **Moderate-to-severe** | 30 | 1994 | 1.5 (0.9, 2.4) | 0.4 (-0.5, 1.2) |
| **Growth stunting<sup>¶</sup>** |  |  |  |  |
| **Moderate-to-severe** | 34 | 2564 | 1.3 (0.9, 2.0) | Referent |
| **No stunting** | 36 | 2994 | 1.2 (0.8, 2.0) | -0.1 (-0.9, 0.6) |
| **DPT vaccination** |  |  |  |  |
| **No doses received** | 23 | 1034 | 2.2 (1.3, 3.9) | Referent |
| **Series completed** | 31 | 3057 | 1.0 (0.6, 1.7) | -1.2 (-2.6, 0.1) |
| **Series incomplete** | 6 | 949 | 0.6 (0.3, 1.3) | -1.7 (-3.0, -0.3) |
| **Injections received in last 12 months** |  |  |  |  |
| **1-12** | 14 | 1436 | 1.0 (0.5, 1.8) | -0.3 (-1.0, 0.5) |
| **13-24** | 3 | 222 | 1.5 (0.6, 3.9) | 0.3 (-1.3, 1.8) |
| **25+** | 3 | 141 | 1.9 (0.5, 7.5) | 0.7 (-2.0, 3.4) |
| **None** | 40 | 3254 | 1.2 (0.8, 2.0) | Referent |
| **Physical violence toward women justified in the household<sup>#</sup>** |  |  |  |  |
| **Justified** | 52 | 3955 | 1.3 (0.9, 2.0) | Referent |
| **Never justified** | 8 | 1100 | 0.7 (0.4, 1.4) | -0.6 (-1.3, 0.1) |

\* Weighted by DHS sampling weights and inverse propensity score weights accounting for sample missingness/exhaustion.
<sup>†</sup> n = 68 (weighted) had indeterminant tetanus serology and were excluded.
<sup>‡</sup> n = 5 (weighted) had missing anemia testing.
<sup>¶</sup> Stunting was assessed using standardized height-for-age scores, with a score of <2 standard deviations indicative of moderate-to-severe stunting. n = 208 (weighted) has missing anthropometric measurements.
<sup>#</sup> A “yes” response by the child’s caretaker to any of the following five questions asking if beating one’s wife is justified if: wife goes out without telling husband; wife neglects children; wife argues with husband; wife refuses sex with husband; wife burns food. These questions are separate from the DHS Domestic Violence Module, which was asked to a different subset of DHS households.

Children with detectable tetanus antibodies had 0.8 (0.1, 1.6) fewer HBsAg-positive cases per 100 children compared with those without detectable antibodies. DPT vaccination reported by caretakers was associated with lower HBsAg-positivity, with children reported to have initiated the series (1-2 doses) having 1.7 (0.3, 3.0) fewer infections per 100 compared with those who had not received any doses (**Table 2**, **Figure 2**). Children in households with reported justification of physical violence toward women had 0.6 (−0.1, 1.3) more HBsAg-positive cases per 100 children compared to children in households where violence was not reported as justified. We observed no significant association between HBsAg-positivity and anemia, growth stunting, or receiving injections in the last 12 months.

### Spatial variation in HBsAg-positivity prevalence, overall and by covariates

Thirty DHS clusters, representing 750-900 households, had an HBsAg-positivity prevalence greater than 10% (**Figure 3**). We observed substantial provincial variation, from 0% (95% CI: 0%, 0%) in several provinces to 5.6% (2.6%, 11.8%) in the northwestern Sud-Ubangi province (**Figure 3, Supplementary Table 3**). Most (21 of 26) provinces had at least one HBsAg-positive child. Twelve provinces had higher HBsAg-positivity prevalence among boys compared with girls, with the largest differences in Sud-Ubangi, Équateur, and Ituri; two provinces (Tshuapa and Kwilu) had higher HBsAg-positivity prevalence among girls compared with boys (**Figure 4, Supplementary Figure 2**). In 10 provinces, children who were tetanus antibody-negative had higher HBsAg-positivity compared with children who were seropositive, with the largest magnitude in Sud-Ubangi (PD: 5.9 per 100 children, 95% CI: 1.4, 10.5) (**Figure 4, Supplementary Figure 3**). Reported DPT vaccination resulted in similar directionality of association as that observed between tetanus seropositivity and HBsAg status in most provinces, but two provinces (both in the Katanga region) had an opposite result, with reporting of any DPT doses associated with higher HBsAg-positivity prevalence (**Supplementary Figure 4**). In no province did HBsAg positivity consistently decrease with decreasing age cohort (**Supplementary Figure 5**).

**Figure 3.**
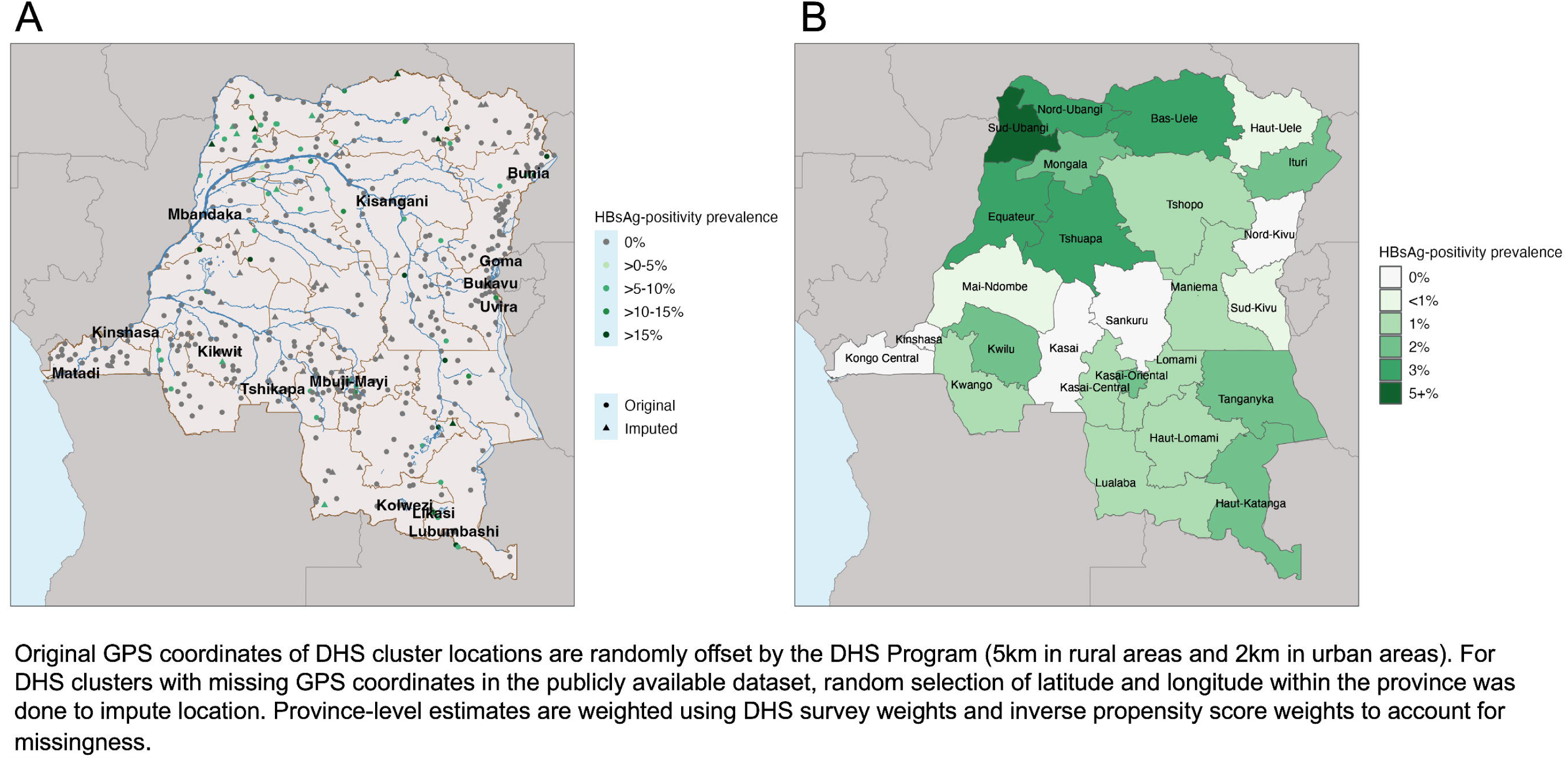
Geographic distribution of the prevalence of HBsAg-positivity in children 6-59 months in the DRC, by **A)** DHS cluster location, with major cities and rivers indicated, and **B)** province.

**Figure 4.**
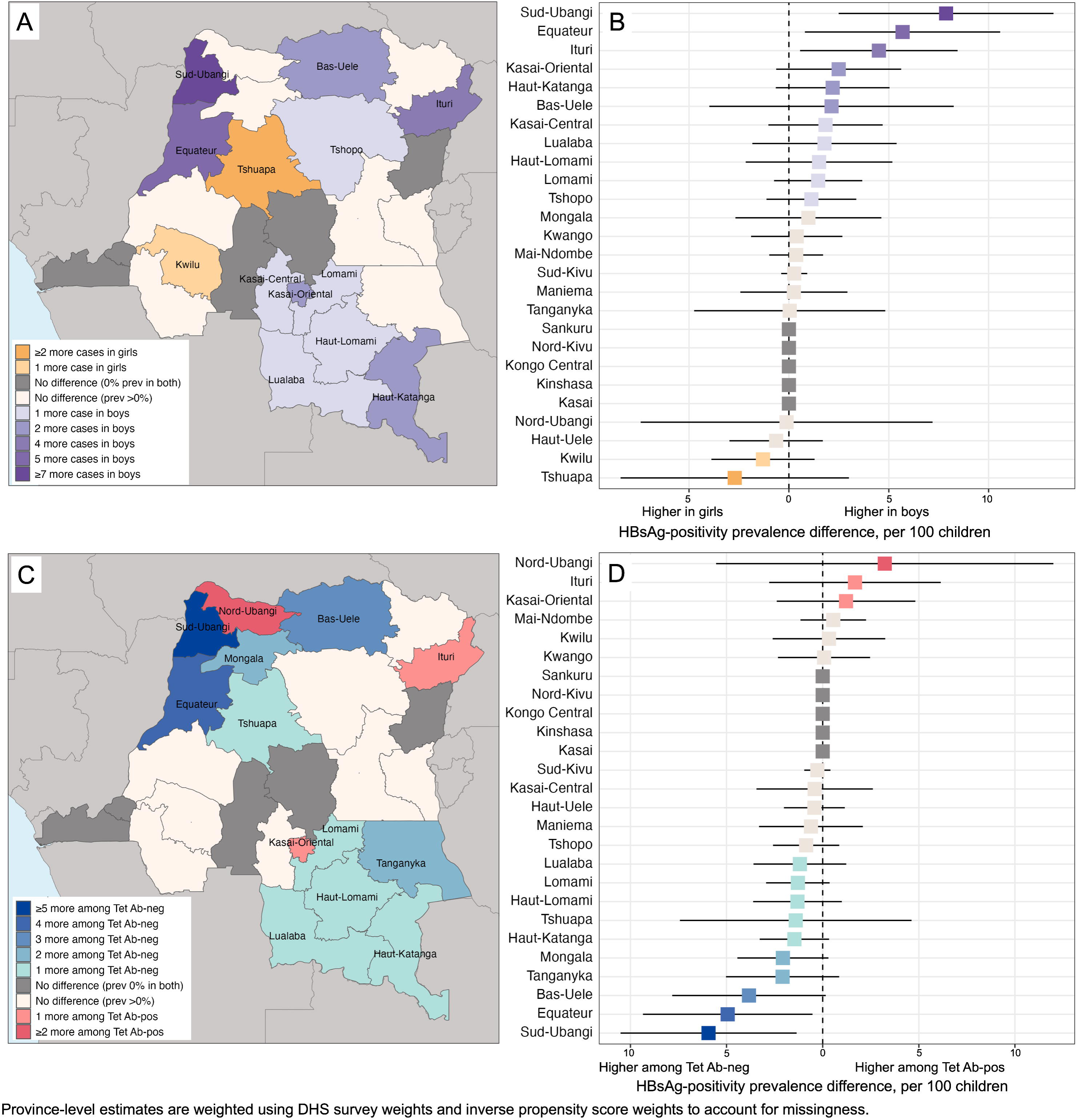
Province-level prevalence differences in HBsAg-positivity in children 6-59 months, by sex (A-B) and by tetanus antibody status (C-D).

### Nested case-control study

In our nested case-control study of children who resided with no HBsAg-positive children (or *other* HBsAg-positive children, for cases), we observed that children living with any HBsAg-positive adult household member had 2.3 (95% CI: 0.7, 7.8) times the odds of HBsAg-positivity compared with children without any HBsAg-positive adult household member (**Table 3**). In particular, children living with a mother who was HBsAg-positive had 7.2 (95% CI: 1.6, 32.2) times the odds of HBsAg-positivity compared to those living with an HBsAg-negative mother.

**Table 3.** Nested case-control study evaluating exposure to an HBsAg-infected adult household member.

| **Exposure** | **Level** | **HBsAg-positive,**<br>**n (%)<sup>\*</sup>** | **HBsAg-negative,**<br>**n (%)<sup>\*</sup>** | **Total<sup>\*</sup>** | **Unadjusted odds**<br>**ratio**<br>**(95% CI)<sup>\*</sup>** |
| --- | --- | --- | --- | --- | --- |
| **HBsAg-positivity of any adult household member** | **≥1 HBsAg-positive adult household member** | 20 (34) | 103 (18) | 123 | 2.3 (0.7, 7.8) |
|  | **No HBsAg-positive adult household members** | 38 (66) | 468 (82) | 506 | Referent |
| **HBsAg-positivity of mother** | **Mother is HBsAg+** | 15 (26) | 27 (5) | 40 | 7.2 (1.6, 32.2) |
|  | **Mother is HBsAg-** | 36 (62) | 460 (84) | 478 | Referent |
|  | **Information about mother unavailable** | 7 (12) | 59 (11) | 64 | -- |

<sup>\*</sup> Weighted by DHS sampling weights and inverse propensity score weights accounting for sample missingness/exhaustion.

## Discussion

We conducted the largest evaluation of HBV epidemiology among children in the DRC to date, using the most recent nationally representative survey and samples from >5,600 children. We found high HBV prevalence (>5-10%) among children aged 6-59 months in specific subnational regions, and an overall national estimate (1.3%) that falls within a “low” range among endemic countries.(31) We further observed that HBsAg-positivity in boys was double that in girls, that HBsAg-positivity prevalence was elevated among children living in rural areas and with less wealth, and that HBsAg-positivity did not decline with decreasing age, as would be expected with improved vaccine distribution over time. These findings highlight populations for focused interventions and emphasize the importance of including subnational estimates for large, heterogeneous countries like the DRC. Indeed, individual provinces in the DRC are larger than many countries on the continent.

Sexual dimorphism in HBV exposure and disease outcomes has been well-documented,(32) but understanding local causes is important for designing effective interventions. Circumcision with insufficiently sanitized instruments(33) and other traditional practices that are sex-specific (such as female genital mutilation(34)) could contribute to differential infection acquisition risk by sex. These practices vary across such a culturally diverse country, but are not well characterized in national surveys.(35,36) Higher prevalence among boys could also be explained by poorer anti-HBs response from vaccination among boys compared with girls, observed previously in Senegal.(37) HBV serology to evaluate protection (anti-HBs) or past exposure (anti-HBc) remains difficult to study in remote settings due to limitations of assay performance on DBS,(38) and is an area for future development. The explanations hypothesized here for the observed prevalence difference by sex relate to modes of horizontal transmission, underscoring the need to consider prevention of horizontal HBV transmission in this context.(11)

Reactive tetanus antibodies and reported DPT vaccination, proxies for HepB3 vaccination, were both associated with lower HBsAg-positivity among children overall. We observed substantial regional variation in this association, with the protective effect of vaccination against HBsAg-positivity observed in only half of provinces. While tetanus serology and caretaker-reported vaccination are imperfect measurements of HBV vaccine receipt, our results suggest that follow-up evaluation of pentavalent vaccine storage, distribution, and administration in these provinces may be worthwhile, as HepB3 and DPT vaccine coverage have declined in the ten years since this survey was conducted.(39,40)

We did not observe a difference in HBsAg-positivity between *P. falciparum* malaria-infected children and -uninfected children, but our study is not optimally designed to interrogate the mechanisms through which an acute infection like malaria could influence HBV infection. Interactions among malaria, HBV infection, and clinical care remain important to evaluate, particularly as the first licensed malaria vaccines (RTS,S and R21), which contain the HBV surface antigen protein as an adjuvant, will soon be implemented in the DRC.(41) RTS,S vaccination has been shown to boost anti-HBs levels,(42) but the impact on HBV prevalence in the population remains unknown.

Living with an HBsAg-positive adult household member, particularly an HBsAg-positive mother, was associated with a higher odds of HBsAg-positivity in children. Horizontal HBV transmission from household members has been documented in settings across the continent and associated in the DRC with sharing of personal items.(43,44) Family members are also commonly donors for blood transfusions, which are frequently needed for malaria-induced anemia and often occur without sufficient infection screening due to test kit stockouts.(45) Prevention of horizontal transmission through proper screening of blood products prior to transfusions, completion of vaccinations, and sanitation of shared personal objects, remains imperative.

This investigation has several limitations. First, our cross-sectional design cannot confirm directionality or timing of exposure, and causality cannot be determined from our unadjusted and ecological estimates, which could be affected by factors not assessed in this study. Second, these samples were analyzed many years after collection, and it is possible that HBsAg protein degraded with time. Reassuringly, however, our national prevalence estimates are slightly lower but generally consistent with those from earlier, smaller studies.(17) Third, the survey was conducted in 2013-2014 and may not reflect the current HBV infection landscape in the DRC. Nonetheless, this analysis provides the most recent nationally representative estimates available in a setting that has yet to implement additional HBV prevention measures.

In this large investigation of HBV in the DRC, we identify regions that might benefit from improved childhood vaccination delivery strategies and community HBV prevention efforts, such as consistent screening of blood products prior to transfusions, sensitization around sanitation of medical instruments, and provision of booster vaccinations. Further, this investigation highlights the importance of subnational prevalence estimates in large countries like the DRC, where estimates from large regions with high HBV prevalence may be diluted within national estimates. As new guidelines for HBV prevention are introduced,(46) this analysis offers a timely investigation in a country in need of improved HBV control measures.

## Acknowledgements

We thank the DHS participants for providing survey responses and samples for this study. We also acknowledge Meghan Cleinmark, Andi Snyder, and the McClendon Clinical Immunology Laboratory for their support in sample processing oversight. We thank the late Steve Meshnick for his mentorship and contributions in the conceptualization of this study.

## Data sharing

Survey data are publicly available upon request through the DHS Program. Biospecimen data are available through the DHS Program.

## Author Contributions

PThompson, JBP, AT, MMK, SBD, FCL, and MY conceptualized the study. PThompson, JBP, AT, MMK, AM, JM, PTshiamala, and GC coordinated sample and materials procurement, and PThompson, JLS, UD, SB, and CEM analyzed samples. CEM, KAP, JKE, FCL, MY, ME, SBD, JBP, and PThompson analyzed the data. CEM and PThompson wrote the first draft, and all authors approved of the final draft.

