## Supplementary Material for "Characterizing hepatitis B virus infection in children in the Democratic Republic of Congo to inform elimination efforts"

**Methods**

**Data source**

The DHS Program used a two-stage cluster design, stratified by province and rural/urban residence, allowing estimation at national, regional, and residential (rural/urban) levels through sample weighting. First, DHS clusters were selected from population representative enumeration area, and second, a random sample of households was selected in each DHS cluster. In every other household selected (50% of all surveyed), a DBS specimen was collected from children aged 6-59 months living in the household, and from adult household members (women 15-49 years and men 15-59 years). Additional details on DHS survey methodology are available in the 2013-14 DRC DHS final report.^1^

**Specimen processing**

For samples of children, one 6-mm DBS punch was eluted in 400µL of 1 x phosphate-buffered saline + 0.25% Triton X-100 in individual 5mL tubes (Sarstedt AG & Co, Nümbrecht, Germany). For samples of adult household members, 6mm punches were previously processed in DRC into 96-well plates; samples selected for this analysis were placed in individual 5mL tubes (Sarstedt AG & Co, Nümbrecht, Germany) for elution. Samples in the elution buffer were incubated at room temperature for 1 hour, and centrifuged at 10,000 x g for 2 minutes. 300 μL (without DBS filter paper or fibers) of eluted sample were placed in individual Sarstedt 2mL tubes and used for serological testing on the Abbott ARCHITECT.

**Variable coding for children 6-59 months:**

- Age of child in months: reported by the parent or guardian on behalf of the child. In the analysis, age in months was grouped into age in years: 0 years = 6-11 months, 1 year = 12-23 months, 2 years = 24-35 months; 3 years = 36-47 months; 4 years = 48-59 months.
- Sex: reported by parent or guardian with binary options for “male” and “female”.
- Rural/urban: designated by the DHS Program prior to conduct of the study.
- Household wealth: composite variable in quintiles calculated by the DHS Program reflecting a household’s standard of living. Wealth quintiles (richest, richer, middle, poorer, poorest) was used as is in the analysis.
- Household size: number of individuals living in the household reported by the respondent (including those not enrolled in any DHS sample).
- Province: 2015 province designations used in descriptive analyses. Pre-2015 provinces (n=11) were used where a smaller number of categorizations was needed.
- Location: setting of household location, analyzed as original response options: “capital, large city” (province-level), “small city,” “town,” or “countryside.”
- Anemia: assessed by hemoglobin concentration using dried blood spot specimens and categorized by the DHS Program into severity levels. In this analysis, severe and moderate levels were grouped together, and mild and no anemia levels were grouped together.
- *Plasmodium falciparum* status: *P. falciparum* infection was determined by quantitative real-time PCR targeting the *P. falciparum* lactate dehydrogenase (*pfldh*) gene, completed by University of North Carolina and previously described. Results have been made available through the DHS program.^2^
- Tetanus antibody: ELISA assay completed by UCLA-DRC program with qualitative results (reactive, nonreactive, indeterminate) available through the DHS Program.^3^ Tetanus and HBV vaccination are both given within the pentavalent vaccine, introduced in 2009, and as such, tetanus serology was used to approximate effectiveness of pentavalent vaccination.
- Growth stunting: standardized height-for-age scores (z-scores) used to categorize into moderate (<2 SDs) or severe (<3 SD) vs no stunting (z ≥ -2 SD).^4^
- Relationship to head of household: reported by the parent or guardian for the child. In the analysis, these were grouped into “child of head of household,” “grandchild of head of household,” and “other.” The “other” category included children who were, in relation to the head of household, nieces/nephews, siblings, adopted/fostered, or other undesignated.

A subset of children (88%) living with their primary caretaker were administered the DHS Children’s Recode Module, and the following variables of interest were analyzed:

- Diphtheria-pertussis-tetanus (DPT) vaccination: documentation by vaccine card or parent or guardian report of DPT doses 1, 2, and 3. At the time of the survey, the national infant immunization program is documented to give the DPT vaccine in a combined pentavalent vaccine that includes hepatitis B. DPT vaccination was analyzed in three ways: number of doses given (0, 1, 2, and 3); complete (3 doses) vs incomplete (1 or 2 doses) vs series not initiated (0 doses); any doses vs no doses.
- Physical violence towards women justified in household: caretakers were asked five questions about whether beating of one’s wife is justified in the household (if wife goes out without telling husband; if wife neglects children; if wife argues with husband; if wife refuses sex with husband; or if wife burns food). In the analysis, these were dichotomized into any “yes” vs “no” to all. These questions are separate from the DHS Domestic Violence Module, which was asked to a different subset of DHS households.
- Injections received: number of injections received in the last 12 months. Categorized into none, 1-12, 13-24, and ≥25 divided to approximate injections per month.
- Use of new, unopened syringe/needle for injections: use of a needle or syringe from an unopened package the last time the child received an injection from a health worker. Analysis subset focused on those answering “yes” or “no,” while “don’t know” or missing are excluded.

**Propensity score weights**

Variables included in the propensity score analysis, based on association with either the outcome (HBsAg status) or DBS sample missingness, were: sex (male, female), age (6-11, 12-23, 24-35, 36-47, 48-59 months), province (list of 26), household setting (provincial capital, small city, town, countryside), household wealth (quintiles from DHS), PCR-confirmed *Plasmodium falciparum* infection (positive, negative/no result), child’s relationship to the head of household, and having slept in the household the previous night. Analytical code in R for this process (as well as the rest of the analysis) is available at: <https://github.com/IDEELResearch/dhs_hbv>.

**Spatial analysis**

Observations with missing GPS were imputed within the province boundary (separate variable available for all) using *st_sample()* from the *sf* package in R.

**Results**

**Supplementary Table 1.** Comparison of sampled children 6-59 months and sampled children 6-59 months with an HBV result (see main Figure 1). Counts are weighted using DHS survey weights (propensity score weights not included).

|  | **Population of 8,547 children 6-59 months, n (%)** | **Population of 5,679 children 6-59 months with HBV results, n (%)** |
| --- | --- | --- |
| **Age (months), mean (SD)** | 32.3 (0.18) | 32.4 (0.22) |
| **Household size, mean (SD)** | 7.0 (0.07) | 6.9 (0.09) |
| **Number of children <5, mean (SD)** | 2.3 (0.03) | 2.2 (0.03) |
| **Age (months), n (%)** |  |  |
| **6-11** | 993 (11) | 625 (11) |
| **12-23** | 1883 (22) | 1261 (22) |
| **24-25** | 1974 (23) | 1284 (22) |
| **36-47** | 1913 (22) | 1278 (22) |
| **48-59** | 1940 (22) | 1280 (22) |
| **Male sex, n (%)** | 4372 (50) | 2900 (51) |
| **Relationship to head of household** |  |  |
| **Child** | 6810 (78) | 4505 (79) |
| **Grandchild** | 1332 (15) | 883 (15) |
| **Other** | 560 (6) | 340 (6) |
| **Rural location, n (%)** | 6071 (70) | 4060 (71) |
| **Location of residence** |  |  |
| **Countryside** | 6071 (70) | 4060 (71) |
| **Provincial capital** | 1494 (17) | 951 (17) |
| **Town** | 926 (11) | 589 (10) |
| **Small city** | 210 (2) | 128 (2) |
| **Wealth, n (%)** |  |  |
| **Poorest** | 1950 (22) | 1288 (22) |
| **Poorer** | 1982 (23) | 1346 (23) |
| **Middle** | 1744 (20) | 1126 (20) |
| **Richer** | 1622 (19) | 1070 (19) |
| **Richest** | 1404 (16) | 897 (16) |
| ***P. falciparum* infection**^†^**, n (%)** | 2504 (34) | 1691 (30) |
| **Moderate-to-severe stunting, n (%)** | 1984 (23) | 2547 (44) |
| **Moderate-to-severe anemia, n (%)** | 2930 (34) | 2010 (35) |
| **Reactive tetanus serology**‡**, n (%)** | 2960 (36) | 2049 (36) |
| **Diphtheria-pertussis-tetanus vaccination**¶ |  |  |
| **Series completed** | 4574 (53) | 2993 (52) |
| **Series incomplete** | 1495 (17) | 955 (17) |
| **No doses received** | 1524 (18) | 1064 (19) |
| **Not available** | 1108 (13) | 716 (12) |
| **Injectable medications received in last 12 months** |  |  |
| **1-12** | 2162 (25) | 1424 (25) |
| **13-24** | 336 (4) | 227 (4) |
| **≥25** | 206 (2) | 143 (2) |
| **None** | 4906 (56) | 3231 (56) |
| **Not available** | 1088 (13) | 701 (12) |
| **Reuse of needles/syringes**# |  |  |
| **New, unopened syringe/needle applied** | 2479 (93) | 1639 (92) |
| **Used/opened syringe/needle applied** | 184 (7) | 127 (7) |
| **Physical violence toward wife reported to be justified in household**** | 5879 (68) | 3955 (69) |

**Supplementary Table 2**. Sensitivity analyses for overall HBsAg-positivity prevalence (weighted)

|  | **HBsAg-positive, n** | **Total, n** | **HBsAg-positivity prevalence per 100 children (95% CI)** |
| --- | --- | --- | --- |
| **S/CO 1** | 98 | 5773 | 1.7 (1.3, 2.3) |
| **S/CO 2** | 81 | 5773 | 1.4 (1.0, 1.9) |
| **S/CO 5 (main)** | 73 | 5773 | 1.3 (0.9, 1.7) |
| **S/CO 100** | 67 | 5773 | 1.2 (0.1, 1.7) |

**Supplementary Table 3. Weighted provincial estimates** for HBsAg-positivity prevalence and 95% confidence intervals for children 6-59 months, corresponding to estimates shown in Figure 2B of the main text.

| **Province** | **HBsAg-positive cases** | **Total sampled** | **HBsAg-positivity prevalence (95% CI)** |
| --- | --- | --- | --- |
| Sud-Ubangi | 17 | 299 | 5.6 (2.6, 11.8) |
| Equateur | 6 | 185 | 3.2 (1.1, 8.9) |
| Bas-Uele | 3 | 109 | 3.1 (0.7, 12.9) |
| Tshuapa | 4 | 129 | 2.9 (1.2, 6.6) |
| Nord-Ubangi | 2 | 82 | 2.9 (1.3, 6.7) |
| Ituri | 5 | 197 | 2.4 (0.7, 7.6) |
| Kwilu | 9 | 443 | 2.0 (0.6, 6.7) |
| Tanganyka | 2 | 112 | 1.7 (0.3, 7.8) |
| Kasai-Oriental | 4 | 231 | 1.6 (0.4, 5.7) |
| Haut-Katanga | 4 | 271 | 1.6 (0.5, 4.5) |
| Mongala | 3 | 186 | 1.5 (0.4, 5.7) |
| Kasai-Central | 3 | 229 | 1.3 (0.4, 4.3) |
| Maniema | 2 | 218 | 1.0 (0.3, 3.6) |
| Kwango | 2 | 267 | 0.9 (0.2, 3.3) |
| Haut-Lomami | 1 | 130 | 0.9 (0.2, 4.1) |
| Lualaba | 1 | 98 | 0.9 (0.1, 7.6) |
| Lomami | 2 | 285 | 0.8 (0.1, 4.9) |
| Tshopo | 1 | 148 | 0.6 (0.1, 4.3) |
| Mai-Ndombe | 1 | 257 | 0.3 (0, 1.9) |
| Haut-Uele | 0 | 93 | 0.3 (0, 2.8) |
| Sud-Kivu | 1 | 428 | 0.1 (0, 1.4) |
| Kinshasa | 0 | 384 | 0 (0, 0) |
| Kongo Central | 0 | 253 | 0 (0, 0) |
| Kasai | 0 | 197 | 0 (0, 0) |
| Sankuru | 0 | 118 | 0 (0, 0) |
| Nord-Kivu | 0 | 421 | 0 (0, 0) |

**Supplementary Figure 1. Sensitivity analyses using different HBsAg S/CO positivity thresholds** for estimated HBsAg-positivity prevalence by province.

S/CO ≥5 was used in this analysis and visualized in Figure 2B of the main text.

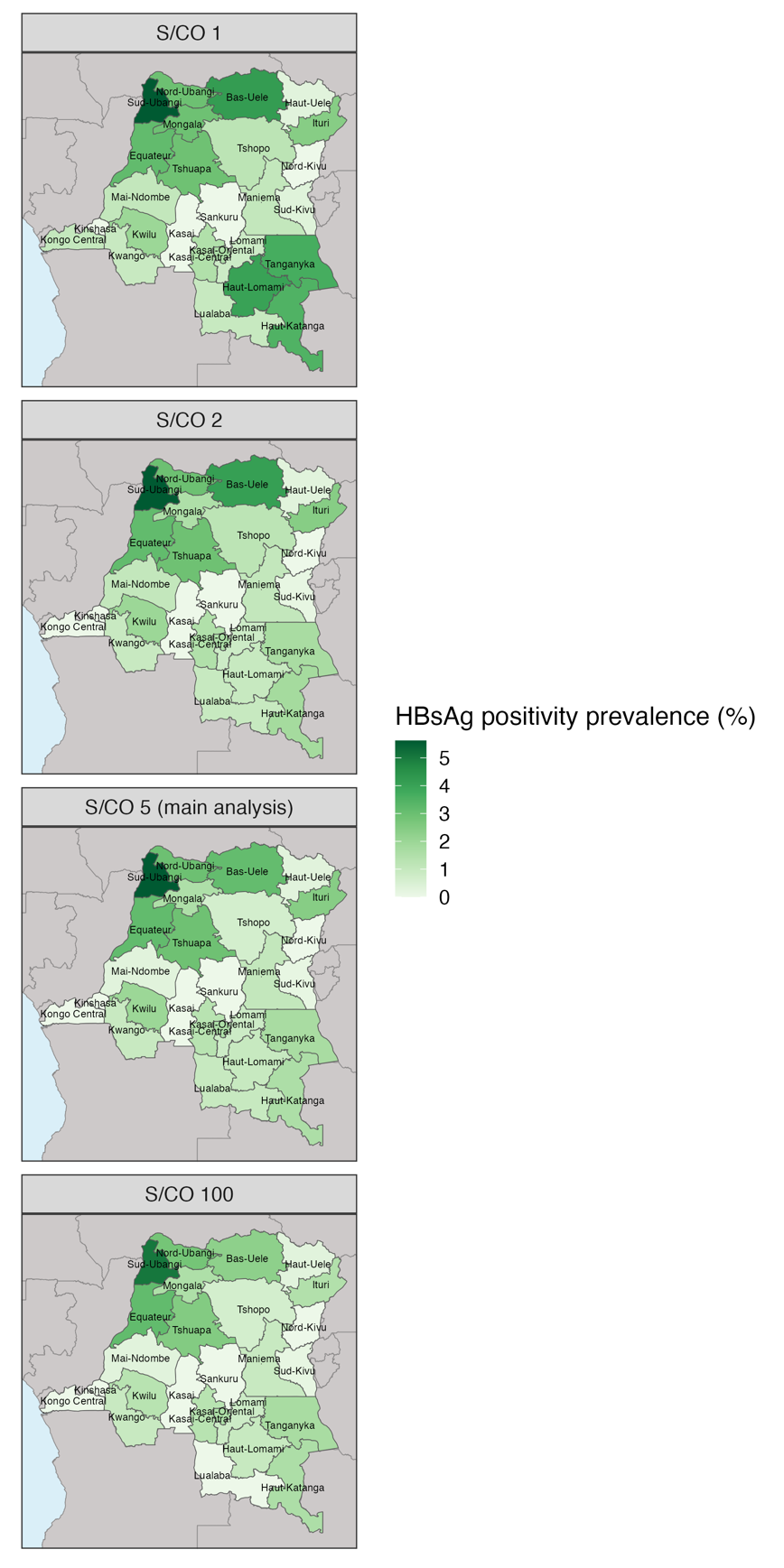

**Supplementary Figure 2. HBsAg-positivity prevalence by sex and province.**

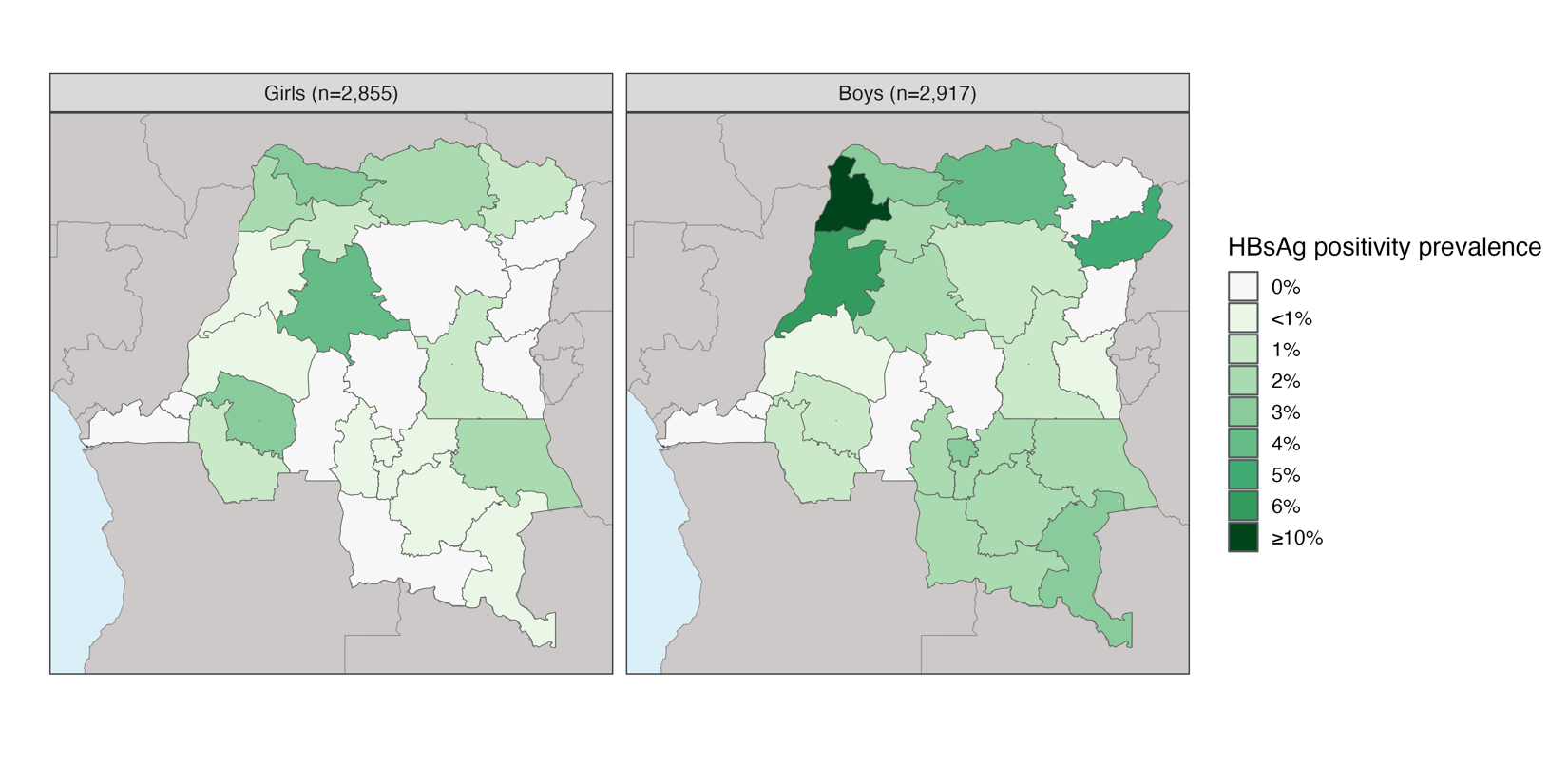

This figure corresponds to prevalence differences by sex in Figure 3A-B in the main text.

**Supplementary Figure 3. Weighted province-level HBsAg-positivity prevalence by nonreactive and reactive tetanus serology.**
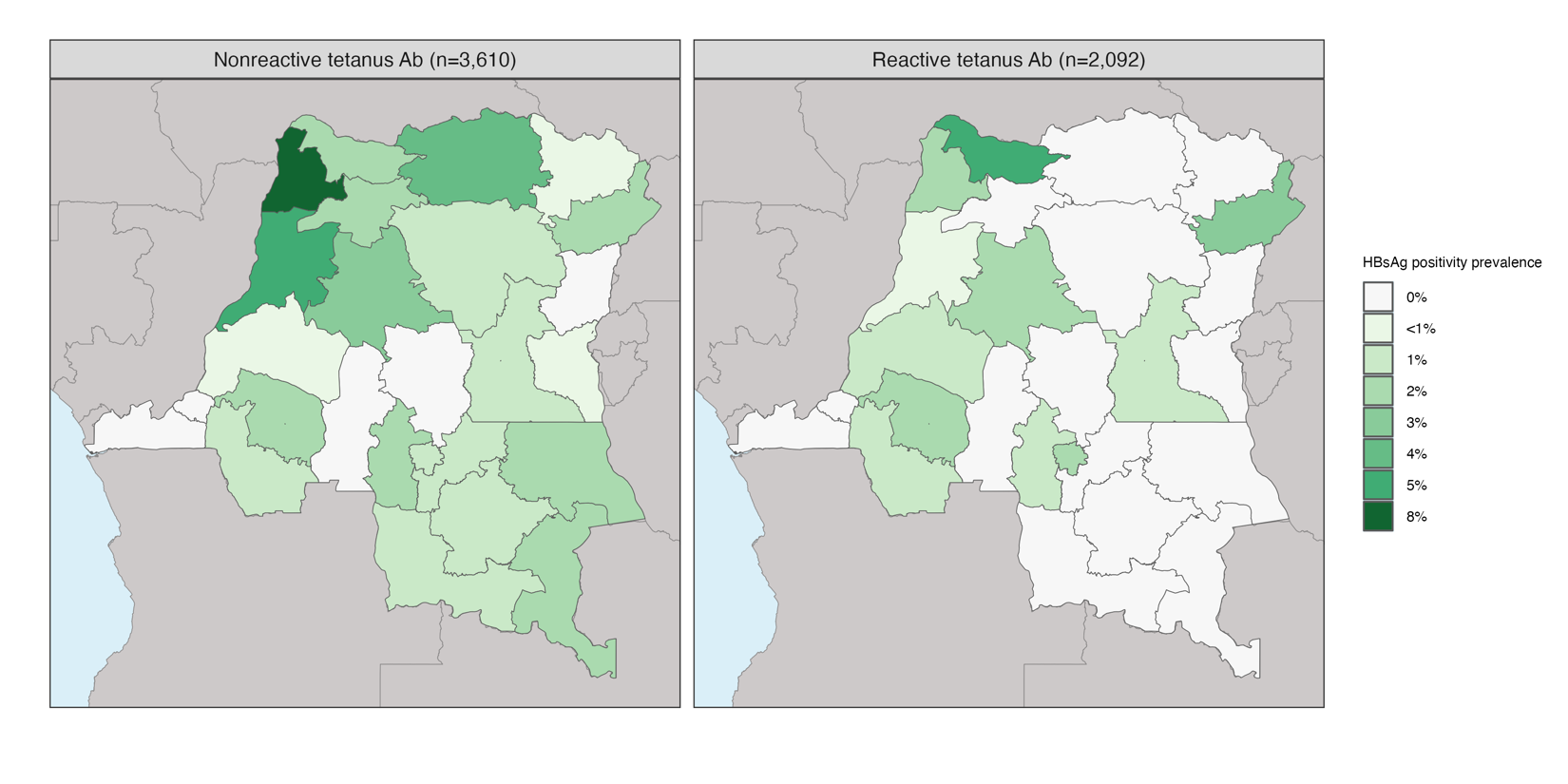

This figure corresponds to prevalence differences shown in Figure 3C-D of the main text.

**Supplementary Figure 4. Weighted province-level HBsAg-positivity prevalence by number of reported DPT vaccine doses**, an alternate to tetanus serology for approximating infant HBV vaccination.

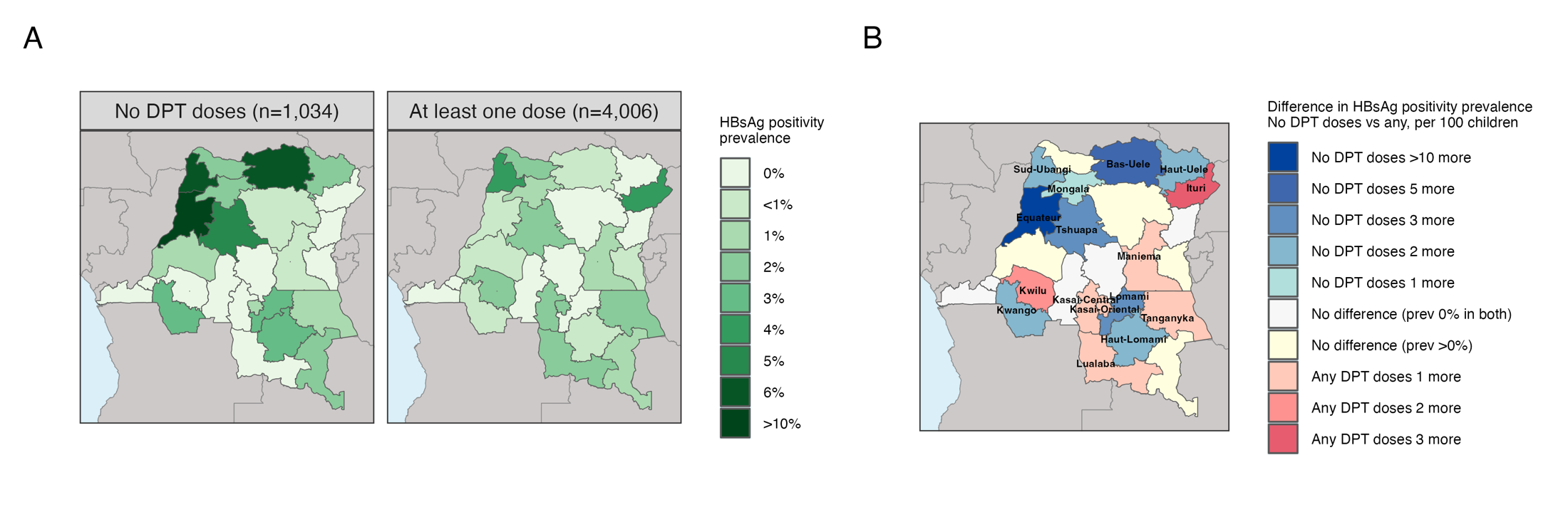

**Supplementary Figure 5. Weighted HBsAg-positivity prevalence by province by child age**, oldest to youngest, mapped (A) and plotted using rolling averages of three-month age groupings (B). In B, the 26 provinces are grouped by region using pre-2015 provinces (n=11).

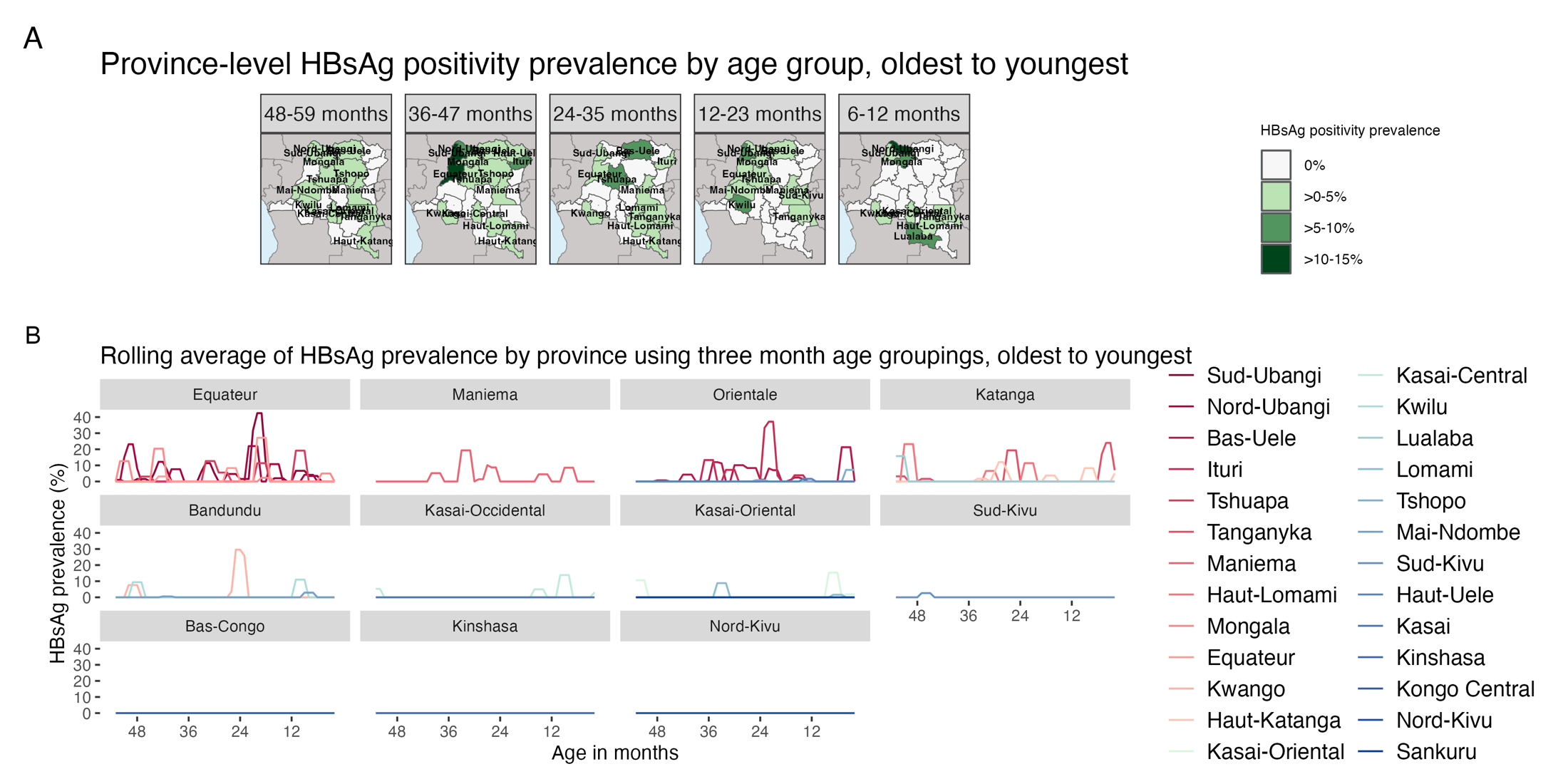
